# Large-scale genome-wide association study to determine the genetic underpinnings of female genital tract polyps

**DOI:** 10.1101/2024.01.29.24301773

**Authors:** Amruta D. S. Pathare, Natàlia Pujol-Gualdo, Valentina Rukins, Jelisaveta Džigurski, Maire Peters, Estonian Biobank Research Team, Reedik Mägi, Andres Salumets, Merli Saare, Triin Laisk

## Abstract

**STUDY QUESTION:** Can a large-scale genome-wide association study (GWAS) meta-analysis identify the genomic risk loci and associated candidate genes for female genital tract (FGT) polyps, provide insights into the mechanism underlying their development, and inform potential overlap with other traits, including endometrial cancer?

**SUMMARY ANSWER:** GWAS meta-analysis of FGT polyps highlighted the potentially shared mechanisms between polyp development and cancerous processes.

**WHAT IS KNOWN ALREADY:** Small-scale candidate gene studies have focused on biological processes such as estrogen stimulation and inflammation to clarify the biology behind FGT polyps. However, the exact mechanism for the development of polyps is still elusive. At the same time, a genome-wide approach, which has become the gold standard in complex disease genetics, has never been used to uncover the genetics of the FGT polyps.

**STUDY DESIGN, SIZE, DURATION:** We performed a genome wide association study (GWAS) meta-analysis including a total of 25,100 women with FGT polyps (International Classification of Disease, ICD-10 diagnosis code N84) and 207,193 female controls (without N84 code) of European ancestry from the FinnGen study (11,092 cases and 94,394 controls) and the Estonian Biobank (EstBB, 14,008 cases and 112,799 controls).

**PARTICIPANTS/MATERIALS, SETTING, METHODS:** A meta-analysis and functional annotation of GWAS signals were performed to identify and prioritise genes in associated loci. To determine associations with other phenotypes, we performed a look-up of associated variants across multiple traits and health conditions, a genetic correlation analysis, and a phenome-wide association study (PheWAS) with ICD10 diagnosis codes.

**MAIN RESULTS AND THE ROLE OF CHANCE:** Our GWAS meta-analysis revealed ten significant (P < 5 x 10^-8^) genomic risk loci. Two signals, rs2277339 (P = 7.6 x 10^-10^) and rs1265005 (P = 1.1 x 10^-9^) (in linkage disequilibrium (LD) with rs805698 r^2^ = 0.75), are exonic missense variants in *PRIM1*, and *COL17A1* genes, respectively. Based on the literature, these genes may play a role in cellular proliferation. Several of the identified genomic loci had previously been linked to endometrial cancer and/or uterine fibroids. Thus, highlighting the potentially shared mechanisms underlying tissue overgrowth and cancerous processes, which may be relevant to the development of polyps. Genetic correlation analysis revealed a negative correlation between sex hormone-binding globulin (SHBG) and the risk of FGT polyps (rg = -0,21, se = 0.04, P = 2.9 x 10^-6^), and on the phenotypic level (PheWAS), the strongest associations were observed with endometriosis, leiomyoma of the uterus and excessive, frequent and irregular menstruation.

**LARGE SCALE DATA:** The complete GWAS summary statistics will be made available after publication through the GWAS Catalogue (https://www.ebi.ac.uk/gwas/).

**LIMITATIONS, REASONS FOR CAUTION:** In this study, we focused broadly on polyps of FGT and did not differentiate between the polyp subtypes. The prevalence of FGT polyps led us to assume that most women included in the study had endometrial polyps. Further study on the expression profile of FGT polyps could complement the GWAS study to substantiate the functional importance of the identified variants.

**WIDER IMPLICATIONS OF THE FINDINGS:** The study findings have the potential to significantly enhance our understanding of the genetic mechanisms involved, paving the way for future functional follow-up, which in turn could improve the diagnosis, risk assessment, and targeted treatment options, since surgery is the only line of treatment available for diagnosed polyps.

**TRIAL REGISTRATION NUMBER:** Not applicable

## Introduction

Polyps of the female genital tract (FGT) are generally benign tissue overgrowths found in both reproductive-aged and postmenopausal women. While the prevalence of polyps can be as high as 50%, most women are asymptomatic. Hence, polyps are usually detected incidentally during routine ultrasound examinations, diagnostic hysteroscopy for other gynaecological disorders, or infertility treatment among women of reproductive age (Hinckley and Milki, 2004; Fatemi *et al*., 2010; Karayalcin *et al*., 2010; Bettocchi *et al*., 2011). However, the occurrence of polyps is age-dependent, with a higher prevalence among postmenopausal women compared to premenopausal women (Dreisler *et al*., 2009). Despite the asymptomatic and benign nature of FGT polyps, for some women, they can negatively impact the daily quality of life by causing abnormal vaginal bleeding and infertility.

Endometrial polyps (EPs) are the most common type of FGT polyps, with a prevalence ranging from 7.8% to 50% (Dreisler *et al*., 2009; de Azevedo *et al*., 2016; Tanos *et al*., 2017). In contrast, endocervical polyps occur only in 2% to 5% of cases, whereas vaginal polyps are rarer (Tanos *et al*., 2017); therefore, limited evidence is available about their biology and clinical implications. Some known risk factors for the development of EPs are advanced age, hypertension, diabetes, obesity, hyperoestrogenism, administration of hormone replacement therapy (HRT), and tamoxifen (Vitale *et al*., 2021; Vieira *et al*., 2022). Nevertheless, the exact pathogenesis of FGT polyps remains unclear. Histologically, EPs are characterised by large, thickened blood vessels with fibrous stroma and irregularly shaped glandular spaces (Tanos *et al*., 2017). Moreover, EPs can be associated with concomitant intrauterine pathologies like endometrial hyperplasia, adenomyosis, endometriosis and chronic endometritis (Annan *et al*., 2012; Raz *et al*., 2021). Despite the benign nature of EPs, around 3.5% of cases progress into carcinoma (Lee *et al*., 2010; Sasaki *et al*., 2018; Uglietti *et al*., 2019). At the same time, the extent of biological mechanisms and pathways shared between EPs and carcinoma, as well as unique molecular characteristics of each condition remain unclear.

Very little is known about the heritability and genetic background of FGT polyps. Genetic factors, including chromosomal translocations in 6p21-22, 12q13-15, or 7q22 regions, may contribute to polypoid morphology (Dal Cin *et al*., 1995; Nijkang *et al*., 2019; Vieira *et al*., 2022), and genetic disorders like Lynch or Cowden syndrome have been reported to be accountable for the development of EPs (Vieira *et al*., 2022). However, a recent study by Sahoo et al. did not confirm the presence of chromosomal rearrangements in endometrial polyps (Sahoo *et al*., 2022).

Thus, the confirmed pathophysiology for the development of FGT polyps is still unknown. However, it can be assumed that the development and mechanism of polyps are multifactorial and can also depend on genetic predisposition. Thus far, the proposed genetic mechanisms are primarily based on small candidate gene studies and are inconclusive (Altaner *et al*., 2006; Pal *et al*., 2008; Banas *et al*., 2018; Doria *et al*., 2018; Takeda *et al*., 2019). Studies in other complex diseases have shown that genome-wide association studies (GWAS) can provide valuable insight into disease biology (Claussnitzer *et al*., 2016), but as far as we are aware, no large-scale GWAS have been published for FGT polyps. Therefore, we performed a GWAS meta-analysis to identify the genetic variants associated with FGT polyps, followed by numerous post-GWAS analyses to understand the shared and unique genetic underpinnings of FGT polyps and endometrial cancer. Ultimately, this knowledge can highlight potential clinical avenues for appropriate diagnosis and management of FGT polyps.

## Materials and methods

### Ethical approval

All Estonian Biobank (EstBB) participants have signed a broad informed consent form, and analyses were carried out under ethical approvals 1.1-12/624 and 1.1-12/2733 from the Estonian Committee on Bioethics and Human Research (Estonian Ministry of Social Affairs) and data release application 6-7/GI/630 from the EstBB. For the FinnGen study, we used only publicly available GWAS summary statistics without individual-level data and thus, a separate ethics approval was not needed.

### Study cohorts

Our analyses included a total of 25,100 women with polyps of the FGT (International Classification of Disease, ICD-10 diagnosis code N84) and 207,193 female controls (without the N84 code) of European ancestry from two studies: summary level statistics from the FinnGen R7 data release (11,092 cases and 94,394 controls) and individual-level data from the EstBB (14,008 cases and 112,799 controls). In FinnGen, cases were defined using the ICD-10 code N84 or corresponding ICD-9 (6210, 6227, 6237, 6246) and ICD-8 (62520) codes. Similar to EstBB, controls were defined as women who did not have the abovementioned disease codes.

To increase study power, we did not distinguish between different types of FGT polyp location in our phenotype definition. However, according to the FinnGen Risteys browser (https://r7.risteys.finngen.fi/phenocode/N14_POLYPFEMGEN), 20% of FGT polyp cases were also cases for phenotype “Uterine polyps”, while in the EstBB data, 70.5 % of N84 cases had a diagnosis for uterine polyps (as defined by the presence of the ICD-10 code N84.0). This most likely reflects differences in the source of phenotype information - FinnGen phenotype definitions mostly use hospital records and thus involve more severe cases, while the EstBB also uses primary care records.

### Cohort-level analyses

The EstBB is a population-based biobank including more than 200,000 individuals (approximately 135,000 of them women) representing 20% of the Estonian adult population. Information on ICD codes is obtained via regular linking with the National Health Insurance Fund and other relevant databases. Individuals with ICD-10 code N84 (mean age at recruitment 49.2 years, standard deviation 12.1) were categorised as cases having been diagnosed with polyps of the genital tract, and all female biobank participants without the diagnosis were considered as controls (mean age at recruitment 44.3 years, sd 16.6). The genotyping procedure for EstBB has been described previously (Koel *et al*., 2023; Laisk *et al*., 2021; Pujol-Gualdo *et al*., 2022). Briefly, Illumina GSAv1.0, GSAv2.0, and GSAv2.0_EST arrays were used for the genotyping of biobank participants at the Core Genotyping Lab of the Institute of Genomics, University of Tartu. Individuals with a call-rate less than 95% and sex mismatch between phenotypic and chromosomal data were excluded from the analysis. Before imputation, genotyped variants were filtered by applying criteria of call rate < 95%, Hardy-Weinberg equilibrium P < 10^−4^ (for autosomal variants only), and minor allele frequency < 1%. Eagle v2.3 software was used for pre-phasing, and Beagle was used for imputation. The population-specific imputation reference of 2297 whole genome sequencing samples was used. Association analysis was performed using REGENIE v2.2.4 with year of birth and 10 principal components as covariates in step I, and variants with a minor allele count < 5 were excluded by default. In downstream association analysis, poorly imputed variants with an INFO score < 0.4 were excluded.

For FinnGen, GWAS summary statistics from the R7 data release were used, and therefore, individual-level data was not available. The summary statistics were obtained from https://www.finngen.fi/en/access_results, whereas the FinnGen cohort and the genotyping/data analysis details have been previously described in Kurki et al. (Kurki *et al*., 2023). To summarise, age, 10 principal components, and genotyping batch were used in REGENIE v2.0.2 analysis as covariates, and for FinnGen summary statistics, variants with a minor allele count > 5 and imputation INFO score > 0.6 were included.

### GWAS meta-analysis

A meta-analysis using fixed-effects inverse variance weighting with genomic control was performed with the GWAMA v2.1 tool. The genome-wide significance level was set to P < 5 × 10^-8^. The genomic inflation factors (lambda) for the individual study summary statistics were 1.046 (EstBB) and 1.045 (FinnGen). Variants present in both cohorts (n = 12,363,169) were included in downstream analyses.

### Annotation of GWAS signals

To annotate the GWAS signals and prioritise potential biologically relevant genes at associated loci, we adopted the following approach.

First, we used the Functional Mapping and Annotation of Genome-Wide Association Studies (FUMA GWAS) platform v1.5.2 to identify genetic association loci. FUMA is an online platform designed for the annotation, prioritisation, and interpretation of GWAS results that uses data from multiple databases to annotate GWAS signals (Watanabe *et al*., 2017). In the first step of this analysis, independent significant variants, lead signals and genomic risk loci were defined. Independent significant variants (IndSigSNPs) were defined as variants that were genome-wide significant (P < 5 x 10^-8^) and had a pairwise LD r^2^ < 0.6, according to the 1000G EUR reference panel. From this subset, lead variants were derived. Finally, risk loci were defined from independent significant SNPs by merging LD blocks if they are less apart than r^2^ < 0.6. Thus, a genomic risk locus can contain several lead SNPs and/or independent significant SNPs, depending on the size of the locus and the LD structure. Thereafter, potential significant candidate SNPs (GWAS meta-analysis P > 0.05) were determined to be in LD with any of the IndSigSNPs (r^2^ ≥ 0.6) within a 1Mb window and had a MAF of ≥ 1%. These candidate SNPs were subjected to further annotation using multiple databases such as Annotate Variation (ANNOVAR) (Wang *et al*., 2010), RegulomeDB scores (ranging from 1 to 7, where lower score indicates greater evidence for having regulatory function) (Boyle *et al*., 2012), and Combined Annotation-Dependent Depletion (CADD) (a continuous score showing how deleterious the SNP is to protein structure/function; scores >12.37 indicate potential pathogenicity) (Kircher *et al*., 2014), 15 chromatin states from the Roadmap Epigenomics Project (ENCODE Project Consortium, 2012; Roadmap Epigenomics Consortium *et al*., 2015), expression quantitative trait loci (eQTL) data (genotype-tissue expression (GTEx) v6 and v7) (GTEx Consortium, 2013) and 3D chromatin interactions from HI-C experiments of 21 tissues/cell types (Schmitt *et al*., 2016). This process provides information on the location, functional impact, and potential regulatory effects of detected SNPs. For eQTL annotations, we focused on tissues similar to uterine/vaginal tissue (GTEx Consortium, 2020).

Using the lead signal identified by FUMA, we performed a look-up in the Open Targets genetics database (Ghoussaini *et al*., 2021). This database combines several layers of evidence across a wide range of cell types and tissues to generate an aggregate ‘variant to gene’ (V2G) score. The V2G score provides identification and correlation of likely causal variants and genes to prioritise the potential functional genes associated with the identified variants. V2G score aggregates several parameters like distance from the canonical transcript start site, eQTLs and protein QTLs (pQTLs) datasets, datasets for chromatin interactions and conformation, molecular phenotypes, and *in silico* functional predictions (using the Variant Effect Predictor or VEP score).

Briefly, we prioritised genes in the identified loci by selecting three genes having the highest V2G score. Then we additionally identified those loci where the GWAS signal includes a missense variant using annotation data from FUMA. To provide additional support for prioritisation and further explore regulatory effects, we looked at eQTL associations according to FUMA annotations, and if none were reported, we looked at potential chromatin interactions as these may also indicate regulatory effects. To gain insight into endometrial-specific eQTLs, we queried the candidate SNPs defined by FUMA in the endometrial eQTL database (Fung *et al*., 2018).

### Gene-based testing

Analysis by Multi-marker Analysis of GenoMic Annotation (MAGMA) v1.6 (de Leeuw *et al*., 2015), with the default settings in FUMA (Watanabe *et al*., 2017), was used to perform gene-based association analysis to complement the single variant analyses. Gene-based analysis enables to detect the joint effect of multiple genetic variants and can thus increase the power to detect associations. Briefly, variants located in the gene body were assigned to protein-coding genes (n = 18,895; Ensembl build 85), and the SNP P-values were merged into a gene test statistic using the SNP-wise mean model (de Leeuw *et al*., 2015). The level of genome-wide significance was set at 0.05/18,895 = 2.6 x 10^-6^, taking into account the number of tested genes.

### Genetic associations with other traits

During the FUMA functional mapping, candidate SNPs were linked with the GWAS catalogue (https://www.ebi.ac.uk/gwas/, GWAS Catalogue e0_r2022-11-29, FUMA v1.5.1) to explore the association of genetic variants with previously published GWAS of different phenotypic traits.

### Genetic correlation analysis

To estimate the genetic correlations between our FGT polyps meta-analysis and 1,335 other traits, we utilised the LD Score Regression method implemented in the Complex Traits Genetics Virtual Lab (CTG-VL) (https://genoma.io/) and 3 additional endometrial cancer traits available in the GWAS catalogue (accession codes GCST006464, GCST006465 and GCST006466) (Bulik-Sullivan *et al*., 2015). Statistical significance was determined by applying a multiple testing correction (FDR < 5%) using the p.adjust function in R v3.6.3.

### Associations with other phenotypic traits

To determine the associations between ICD-10 diagnosis main codes and the N84 diagnosis of polyps in the FGT, we conducted an analysis using individual level data from the EstBB. Logistic regression was used to test the associations between N84 and other ICD-10 codes while controlling for age at recruitment and 10 genetic principal components to account for population stratification and avoid false associations due to ancestry/regional differences between cases and controls. Age was included as a covariate to account for incomplete electronic diagnosis data for older participants in the biobank. Our analysis was limited to the diagnosis main codes along with all the subcodes to increase the power of the data. Statistically significant associations were determined by applying Bonferroni correction (2000 tested ICD main codes, corrected P threshold of 2.5 x 10^-5^). Odds ratios (ORs) were calculated by the logistic regression method and represented as adjusted ORs. The resulted associations were filtered to remove the diagnoses related to exogenous factors (such as injuries, poisoning, accidents, assaults, etc. in the S, T, U, V, W, X, and Y subchapters) and the PheWAS library (https://github.com/PheWAS/PheWAS) was used to visualise the results. All analyses were performed using R v4.1.3

In the FinnGen data, we did not have access to the individual level data, but using the Risteys portal (https://r7.risteys.finngen.fi/phenocode/N14_POLYPFEMGEN), we explored the results of the survival analysis evaluating associations between FGT polyps and other selected phenotypes. A detailed description of the survival analysis can be found in the Risteys documentation (https://r7.risteys.finngen.fi/documentation), but briefly, this type of analysis tests the association between an exposure endpoint and an outcome endpoint. For example, in the context of FGT polyps, what is the association between a diagnosis of FGT polyp (exposure endpoint) and endometrial carcinoma (outcome endpoint).

## Results

### Summary of GWAS

GWAS of a total of 25,100 women with polyps of the FGT and 207,193 female controls revealed ten significant (P < 5 x 10^-8^) genomic risk loci and 23 independent signals (Fig. 1, Table 1). The strongest signal, rs1702136 (P = 2.17 x 10^-19^), was observed on chromosome 3, which is an intronic variant of the *EEFSEC* gene. The majority of the significant signals (91.3%) were either intronic or intergenic variants, whereas two signals, rs2277339 (P = 7.57 x 10^-10^) and rs1265005 (P = 1.09 x 10^-9^, in LD with rs805698, r^2^ = 0.75) on chromosomes 12 and 10, respectively, were exonic missense variants.

**Figure. 1.**
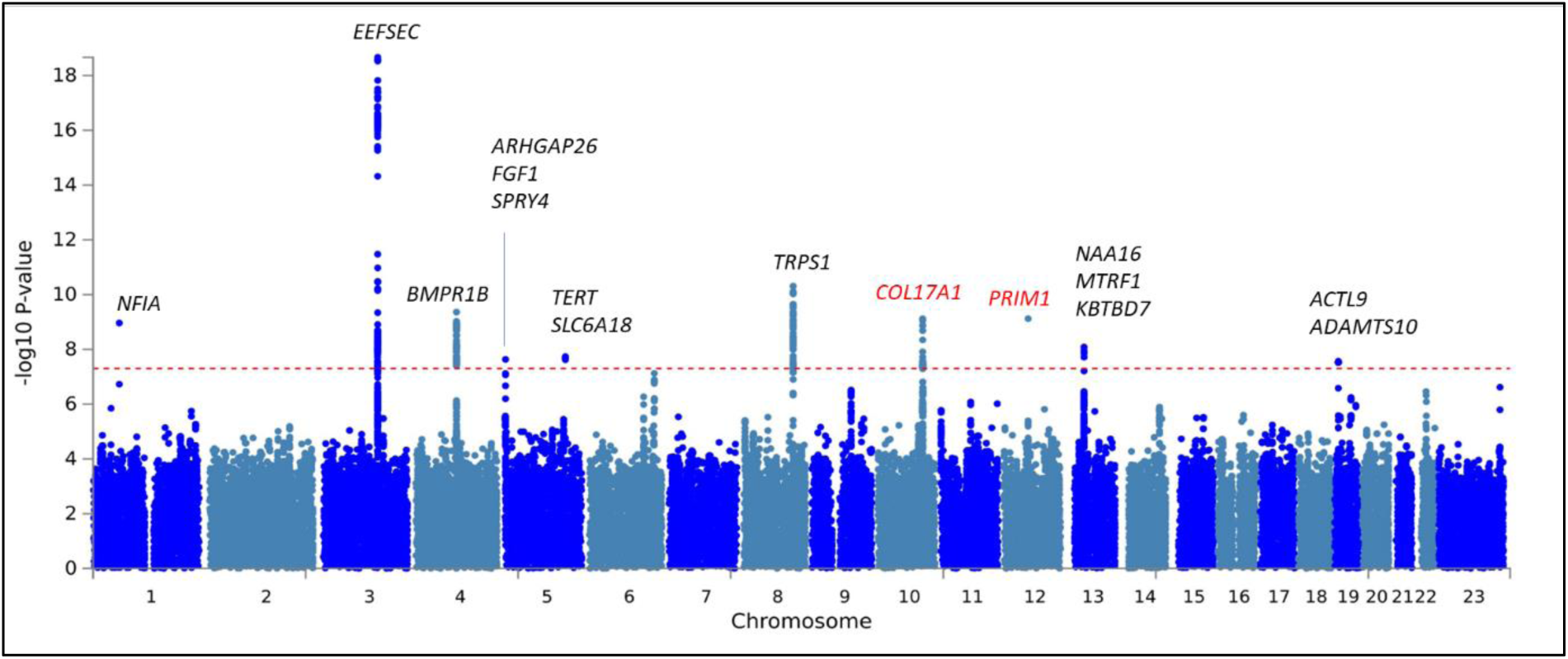
Manhattan plot for significant genomic risk loci identified for polyps of female genital tract. On the Manhattan plot, the x-axis represents chromosomes, while the y-axis represents −log10(P-values) for the association of variants identified in polyps of the female genital tract. The horizontal red dashed line represents the genome-wide significance threshold (P < 5 × 10^−8^). The prioritised genes for each locus are labelled on the top, and genes associated with exonic missense variants are coloured red.

**Table 1.**
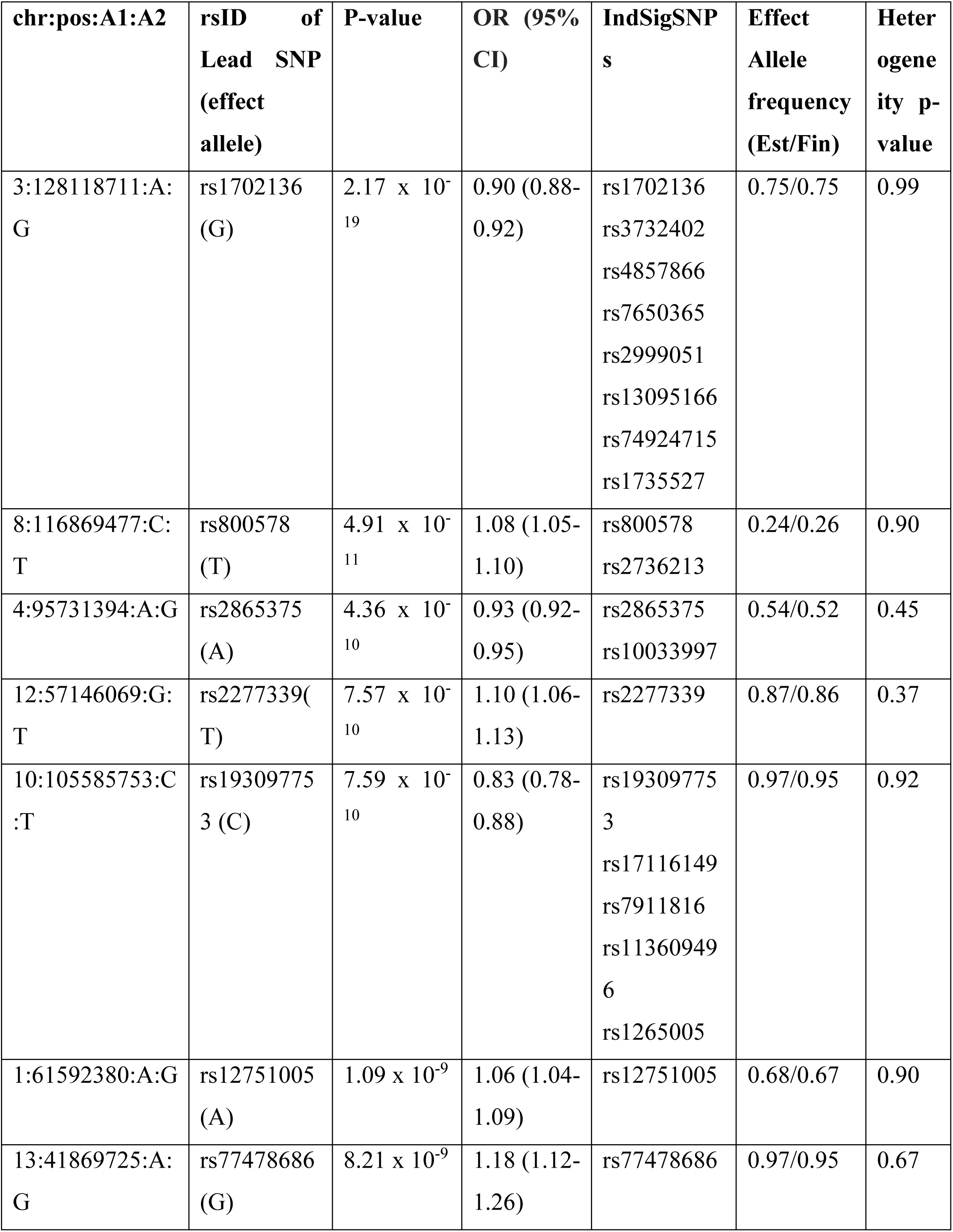
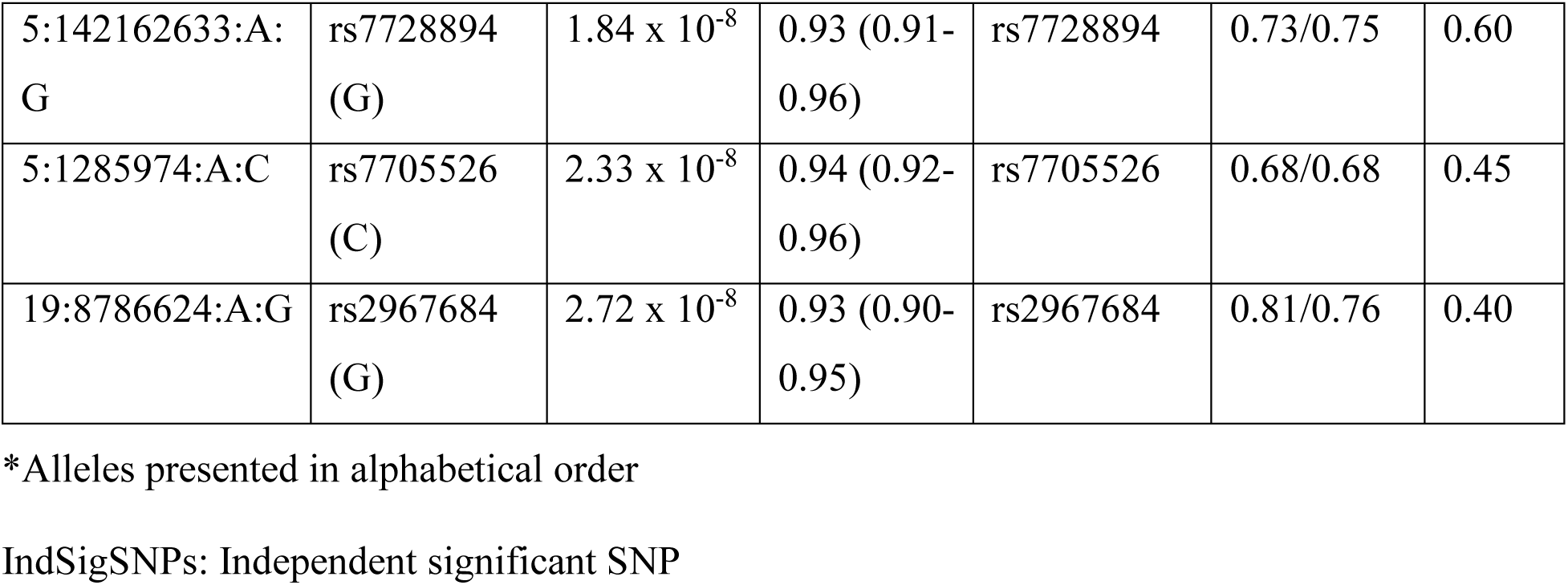
Summary statistics of significant genomic risk loci with lead SNP and independent significant SNP.

### Functional annotation of associated variants and gene prioritisation

We performed a look-up in the Open Targets Genetics database to evaluate the functional association of identified genomic loci and potentially mapped genes reflected by the V2G score. We additionally highlighted those loci where the GWAS signal includes a missense variant using annotation data from FUMA (Table 2). Full details of gene prioritisation together with supporting evidence from FUMA eQTL and chromatin interaction mapping, can be found in Supplementary Table 1. Based on coding variants in our GWAS signals, we were able to prioritise *PRIM1* and *COL17A1* in loci on chromosomes 12 and 10, respectively. PRIM1 is a DNA primase involved in the initiation of DNA replication by synthesising RNA primers for Okazaki fragments during discontinuous DNA replication (Shiratori *et al*., 1995). rs2277339 is also a cis-eQTL for *PRIM1* in endometrial tissue, which further supports *PRIM1* as a candidate gene in this locus. *COL17A1*, on the other hand, is primarily involved in cellular migration, cellular differentiation, and extracellular matrix organisation (Jones *et al*., 2020). While these genes have not yet been directly associated with FGT polyps, their biological functions, particularly those related to cellular proliferation, highlight their potential involvement in the development of polyps.

**Table 2.**
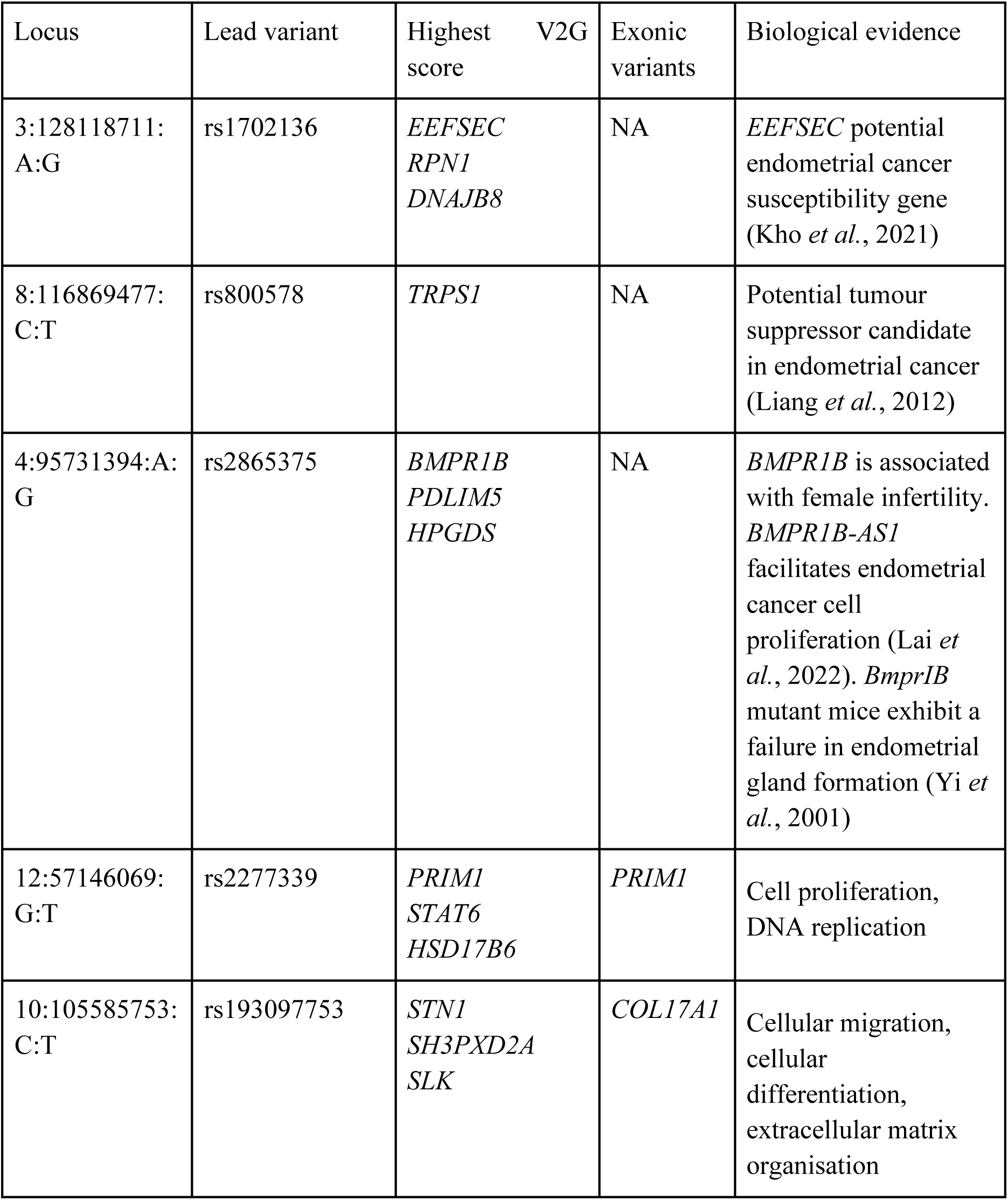
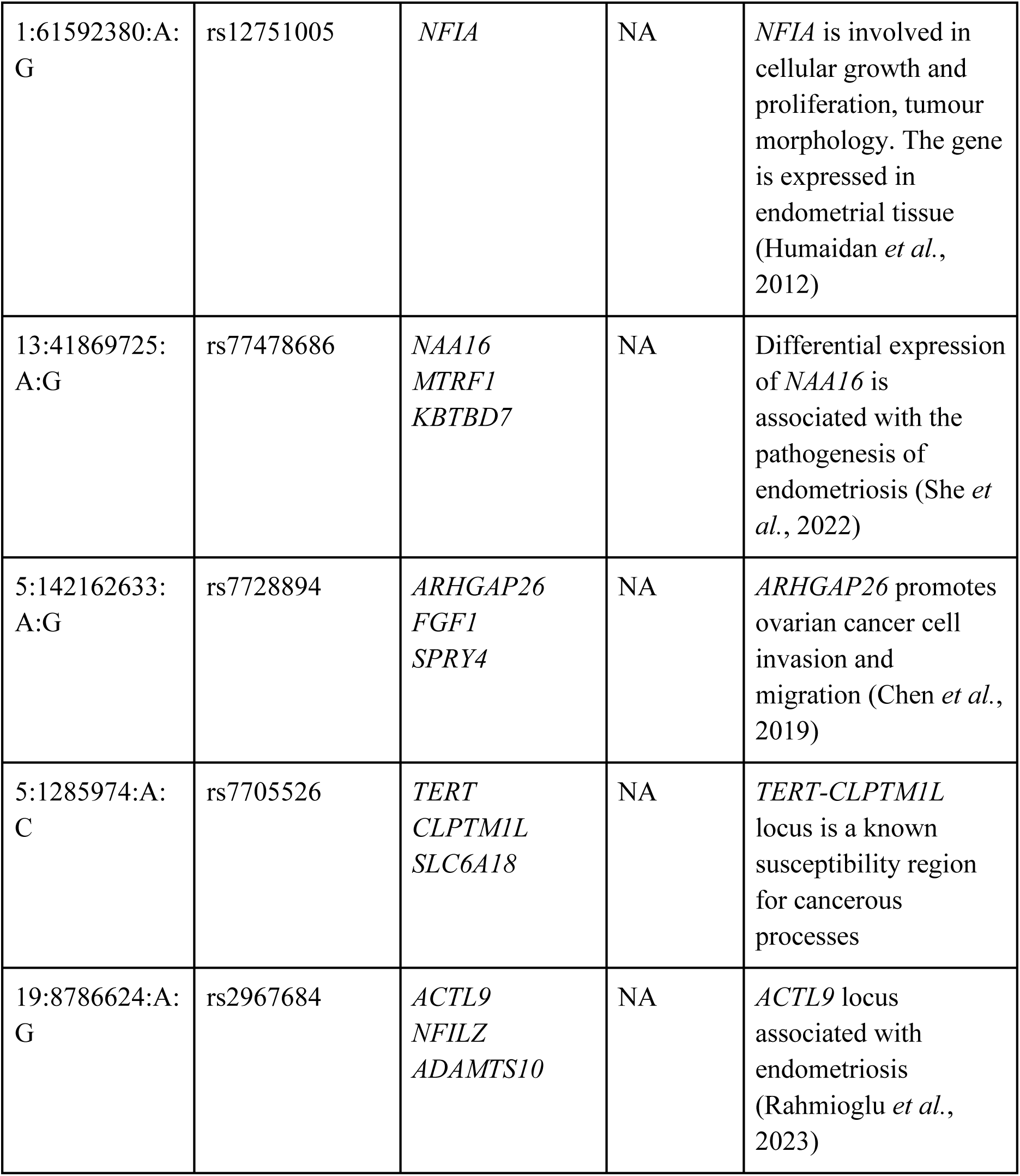
Summary of gene prioritisation results.

| Locus | Lead variant | Highest V2G score | Exonic variants | Biological evidence |
| --- | --- | --- | --- | --- |
| 3:128118711:<br>A:G | rs1702136 | *EEFSEC*<br>*RPN1*<br>*DNAJB8* | NA | *EEFSEC* potential endometrial cancer susceptibility gene (Kho *et al.*, 2021) |
| 8:116869477:<br>C:T | rs800578 | *TRPS1* | NA | Potential tumour suppressor candidate in endometrial cancer (Liang *et al.*, 2012) |
| 4:95731394:A:<br>G | rs2865375 | *BMPR1B*<br>*PDLIM5*<br>*HPGDS* | NA | *BMPR1B* is associated with female infertility. *BMPR1B-AS1* facilitates endometrial cancer cell proliferation (Lai *et al.*, 2022). *Bmpr1B* mutant mice exhibit a failure in endometrial gland formation (Yi *et al.*, 2001) |
| 12:57146069:<br>G:T | rs2277339 | *PRIM1*<br>*STAT6*<br>*HSD17B6* | *PRIM1* | Cell proliferation, DNA replication |
| 10:105585753:<br>C:T | rs193097753 | *STN1*<br>*SH3PXD2A*<br>*SLK* | *COL17A1* | Cellular migration, cellular differentiation, extracellular matrix organisation |
| 1:61592380:A:<br>G | rs12751005 | *NFIA* | NA | *NFIA* is involved in cellular growth and proliferation, tumour morphology. The gene is expressed in endometrial tissue (Humaidan *et al.*, 2012) |
| 13:41869725:<br>A:G | rs77478686 | *NAA16*<br>*MTRF1*<br>*KBTBD7* | NA | Differential expression of *NAA16* is associated with the pathogenesis of endometriosis (She *et al.*, 2022) |
| 5:142162633:<br>A:G | rs7728894 | *ARHGAP26*<br>*FGF1*<br>*SPRY4* | NA | *ARHGAP26* promotes ovarian cancer cell invasion and migration (Chen *et al.*, 2019) |
| 5:1285974:A:<br>C | rs7705526 | *TERT*<br>*CLPTM1L*<br>*SLC6A18* | NA | *TERT-CLPTM1L* locus is a known susceptibility region for cancerous processes |
| 19:8786624:A:<br>G | rs2967684 | *ACTL9*<br>*NFILZ*<br>*ADAMTS10* | NA | *ACTL9* locus associated with endometriosis (Rahmioglu *et al.*, 2023) |

Several of the prioritised candidate genes had previously been associated with (endometrial) cancer - *EEFSEC*, *TRPS1*, *TERT/CLPTM1L*, one could be directly linked with endometrial biology (*BMPR1B*), and for the remaining, their biological significance in FGT polyps remains unclear.

### Gene-based associations of female genital tract polyps

To combine the joint effect of multiple genetic variants and increase the power to detect associations, we performed a MAGMA gene-based test implemented in FUMA. Eight genes were identified which passed the recommended threshold for significance (P = 2.6 × 10^-6^, Bonferroni correction for association testing of 18,895 protein-coding genes): *EEFSEC, TERT, RUVBL1, BMPR1B, TRPS1, COL17A1, BET1L, WBP4* (Supplementary Figure 1, Supplementary Table 2). Majority of these associations mirror the genes prioritised in the single variant analysis, while the *BET1L* locus was not genome-wide significant in the single variant analysis and is thus novel. Previously, *BET1L* has been associated with endometrial cancer (Bateman *et al*., 2017) and uterine fibroids (Cha *et al*., 2011; Edwards *et al*., 2013; Liu *et al*., 2018).

### GWAS catalogue look-up

We searched the GWAS catalogue for associations between previously published phenotypic traits and candidate SNPs identified by the FUMA tool to gain additional insight into their potential biological roles (Fig. 2, Supplementary Table 3). Based on the GWAS catalogue look-up, the *TERT-CLPTM1L* (rs7705526) locus was clearly associated with cancers, including the development of reproductive cancers such as ovarian cancer and prostate cancer. Potential involvement in cancerous processes was also observed for the *EEFSEC* locus (rs4857866), which was associated with prostate cancer and also nominally with endometrial cancer (P < 1 x 10^-6^). Several identified signals were related to reproductive traits, including menarche (rs2277339-*PRIM1*), menopause (rs2277339-*PRIM1*), gestational age (rs4857866-*EEFSEC*) and uterine fibroids (rs2277339-*PRIM1*, rs193097753-*COL17A1*, rs17116149-*COL17A1*). The rs193097753 signal was additionally associated with the known risk factors for the development of FGT polyps, such as type 2 diabetes (P < 5 x 10^-8^) and waist-to-hip ratio (P < 2 x 10^-11^), further substantiating its functional significance.

**Figure. 2:**
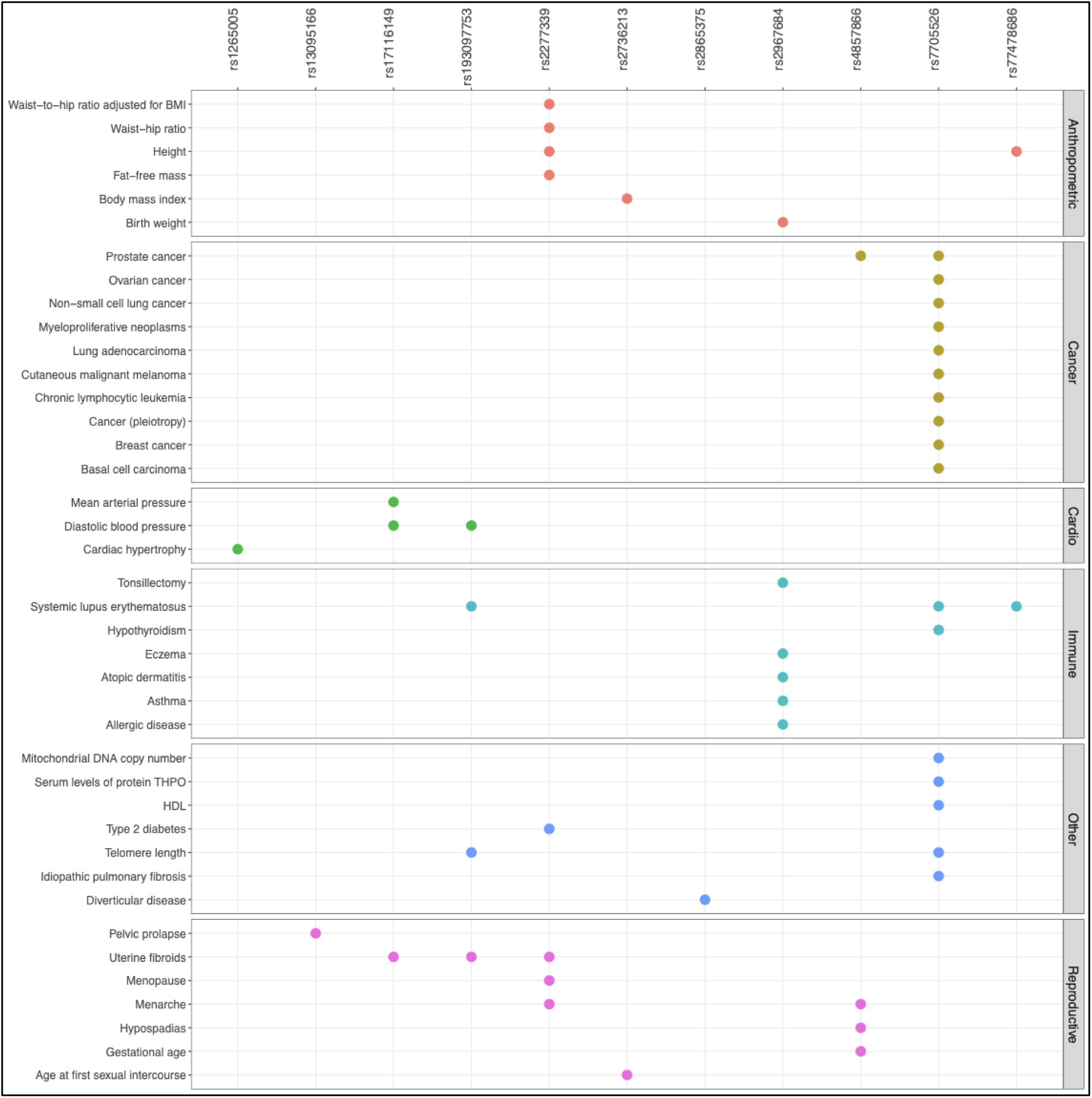
GWAS catalogue look-up showing associations between genetic variants associated with female genital tract polyps and other phenotypes. The figure highlights the genome-wide significant (P < 5 x 10^-8^) association between FGT polyps genetic risk factors (individual significant SNPs) and previously published phenotypic traits and disorders.

### Look-up of variants associated with endometrial cancer

Since there is some overlap between FGT polyp genetic risk loci and those known to be associated with (endometrial) cancer, we conducted a look-up of variants associated with endometrial cancer (O’Mara *et al*., 2018) in our GWAS data. Of the 19 SNPs queried, seven were nominally significant (P < 0.05) (rs9668337-*SSPN*, rs1740828-*SOX4*, rs17601876-*CYP19A1*, rs882380-*SNX11*, rs3184504-*SH2B3*, rs1129506-*EVI2A*, rs11263761-*HNF1B*) in the FGT polyp analysis as well (Supplementary Table 4). When querying the 10 lead variants associated with FGT polyp in the endometrial cancer summary statistics, we observed that three were nominally significant (P < 0.05) in the endometrial cancer and also in the endometrial cancer with endometrioid histology studies (rs1702136-*EEFSEC*, rs2865375-*BMPR1B*, rs7705526-*TERT*).

### Genetic correlation between FGT polyps and other traits

We conducted pairwise genetic correlation (rg) analyses to examine the relationship between polyps of the FGT and 1,335 different traits obtained from the Complex Traits Genetics Virtual Lab (CTG-VL, https://genoma.io/) and 3 additional endometrial cancer traits available in the GWAS catalogue (accession codes GCST006464, GCST006465 and GCST006466). After running the analysis and correcting for multiple testing, we identified a total of 11 significant (FDR < 0.05) genetic correlations. Selected genetic correlations between polyps and various phenotypic categories such as anthropometrics, cardiovascular diseases, genitourinary traits, mood disorders, sex hormones and surgical procedures are displayed in Fig. 3, full results in Supplementary Table 5. As expected, a positive genetic correlation was observed with genitourinary traits and hysterectomy, emphasising that polyps are commonly occurring incidental findings in gynaecological disorders. Furthermore, sex hormone-binding globulin (SHBG) was negatively correlated with the risk of FGT polyps. We were not able to detect a significant genetic correlation between FGT polyps and estradiol level or BMI, known risk factors of FGT polyps, in our dataset. FGT polyps showed weak correlation with two of the three endometrial cancer phenotypes tested (endometrial cancer and endometrial cancer - endometrioid histology, both rg 0.25-0.27, se 0.12-0.13), but these associations were only nominally significant (P = 0.03) and did not pass multiple testing correction threshold.

**Figure 3:**
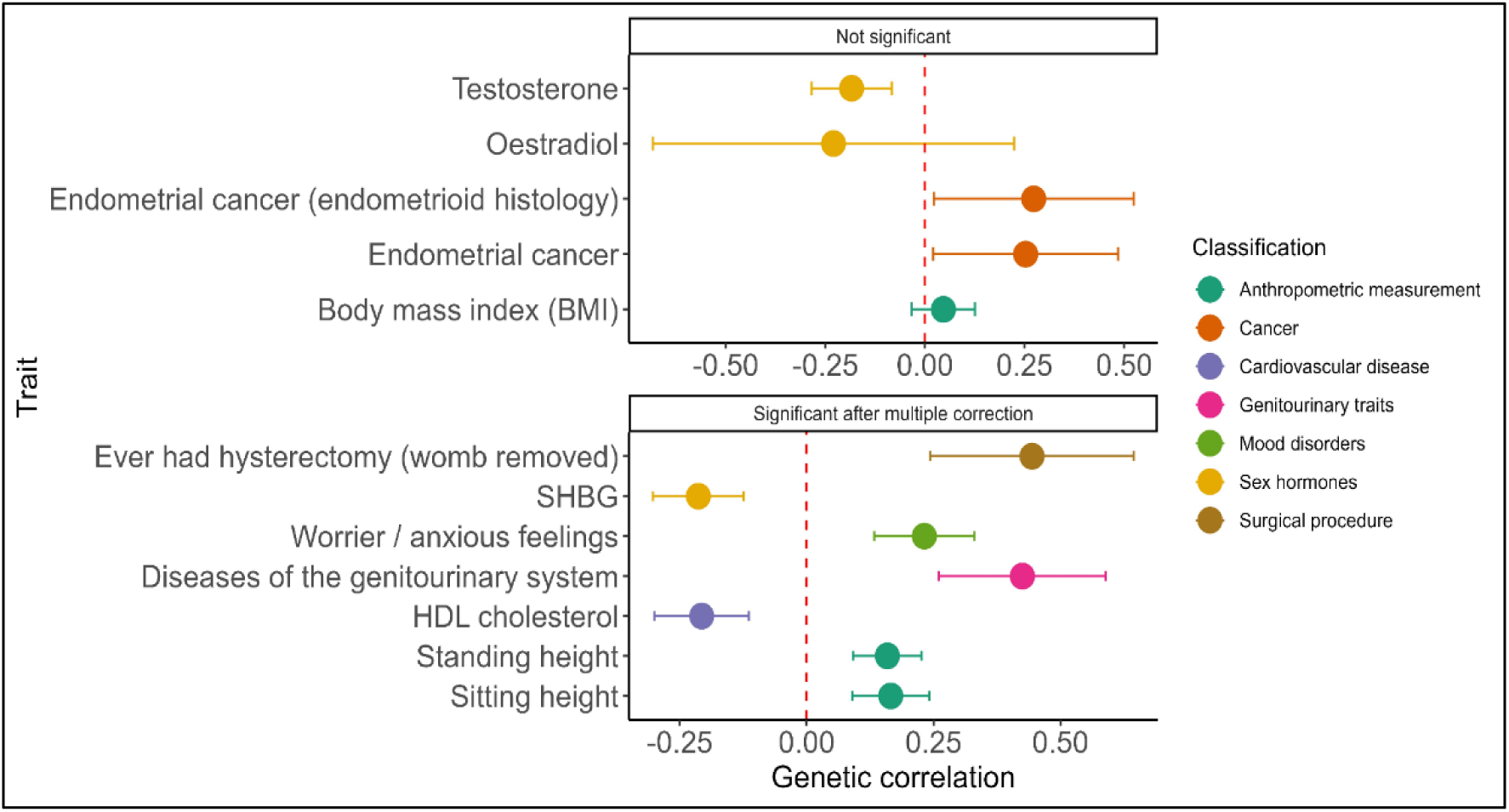
Results of genetic correlation analysis between endometrial polyps and other relevant traits. The plot displays selected genetic correlations between female genital tract polyps and traits from various phenotypic categories such as anthropometrics, cardiovascular traits, genitourinary traits, mood disorders, sex hormones and surgical procedures that are colour-coded. Significant associations after FDR multiple correction are shown in the lower panel, while the upper panel shows selected traits of interest that do not pass the multiple testing correction. The centre dot marks the estimated genetic correlation (rg) value, and error bars indicate 95% confidence limits. The dotted red line indicates no genetic correlation.

### Associations between polyps of FGT and other diagnosis codes

Additionally, to evaluate other phenotypes’ association with FGT polyps, we performed a phenome-wide phenotype association analysis using the EstBB data for disease codes. All 239 phenotypic associations with a P < 2.5 x 10^-5^ (corresponding to a Bonferroni-corrected threshold of 0.05/2020) are shown in Supplementary Table 6. The most significant association of FGT polyps related to other phenotypes were: i) leiomyoma of uterus (D25), which are uterine fibroids known to often occur together with EPs (Kınay *et al*., 2016); ii) endometriosis (N80), which has been correlated with a high prevalence of endometrial polyps; iii) excessive, frequent and irregular menstruation (N92) which is one of the classical symptoms of EPs, and; iv) other noninflammatory disorders of uterus except cervix (N85) (Fig. 4). Moreover, women with a diagnosis for FGT polyps had significantly more diagnoses for benign neoplasms in several tissues, such as the uterus (D26), ovary (D27), unspecified female genital organs (D28 and D39) and skin (D23) as well as malignant neoplasms of the uterus (C54) and skin (C44). In our analysis, nasal polyps (J33) and K62 (which includes anal/rectal polyps) were also nominally significant, which supports the idea of some systemic tissue overgrowth in epithelial tissues similar to endometrial polyps harbouring glandular and luminal epithelium.

**Figure 4:**
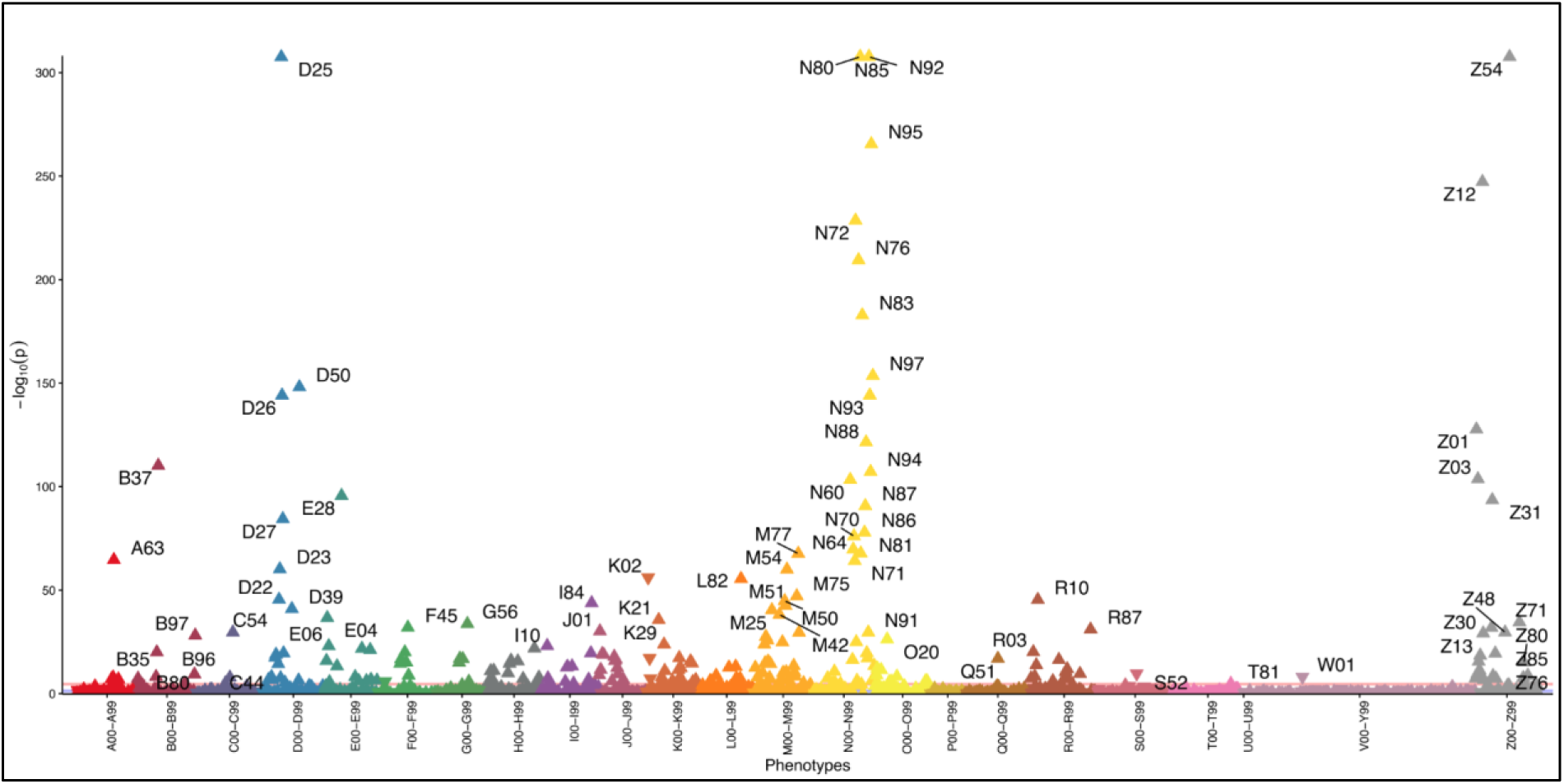
Association of Polyps of female genital tract (N84) with other phenotypes. Each triangle in the plot corresponds to one ICD10 main code, and different colours represent different diagnosis categories. The direction of the triangle represents the direction of effect and upward-pointing triangles show increased significance of diagnosis code in female genital tract polyps. The red line indicates the Bonferroni corrected threshold for statistical significance.

For the highlighted associations, we queried the FinnGen Risteys R7 portal to see if we observe similar associations in the Finnish data. While the analysis in the Estonian data does not consider which diagnosis in the tested pair comes first, the FinnGen data survival analysis tests the association in both directions. Uterine leiomyoma, endometriosis, other benign neoplasm of uterus, benign neoplasm of ovary, malignant neoplasm of uterus, and carcinoma in situ of endometrium were significantly associated with FGT polyps in the survival analysis in FinnGen data, (Supplementary Table 7) both if the diagnosis occurred before, or after the FGT polyp diagnosis. The results of phenotypic associations of FGT polyps are in concordance with the observed genetic associations and point to the fact that benign and cancerous processes share some mechanisms (such as cellular proliferation) on a genetic level.

## Discussion

Gynaecological polyps are a common diagnosis in women, with potential negative implications for women’s reproductive health and well-being. We conducted the first large-scale GWAS meta-analysis to study the genetic underpinnings of FGT polyps and their association with other phenotypic traits from two European ancestry biobanks. The analysis revealed ten genomic risk loci, of which two (rs2277339 and rs193097753) tagged exonic missense variants suggestive of plausible functional importance in the development of polyps. Furthermore, several of the identified genetic risk loci have previously been associated with (endometrial) cancer and/or uterine fibroids. Genetic correlation analysis additionally showed negative correlation with SHBG levels, but no statistically significant genome-wide correlation with endometrial cancer. PheWAS analysis showed an increased prevalence of endometriosis, irregular and excessive menstruation, uterine fibroids and neoplasms of the uterus and other tissues in women with FGT polyps.

Even though most EPs are benign, there is still a certain level of risk these polyps may progress to malignant transformation (Antunes *et al*., 2007; Lenci *et al*., 2014). According to a systematic review and meta-analysis, 2.7%-3.6% of EPs can be considered malignant, with significant heterogeneity in prevalence among pre- and postmenopausal women (Lee *et al*., 2010; Sasaki *et al*., 2018; Uglietti *et al*., 2019). While we saw no statistically significant genetic correlation between FGT polyps and endometrial cancer, several of the mapped genetic risk loci/prioritised genes (rs1702136-*EEFSEC*, rs800578-*TRPS1*, rs2865375-*BMPR1B*, and rs7705526-*TERT-CLPTM1L*) have previously been associated with cancer either in GWAS or functional studies (Liang *et al*., 2012; Wu *et al*., 2014; Kho *et al*., 2021; Dos Santos *et al*., 2022), most likely reflecting the fact that both benign and malignant tissue overgrowths may utilise the same cellular mechanisms to some extent. This is supported by the results of the associated diagnoses analysis where women with a diagnosis of FGT polyp also have more diagnoses related to benign and malignant neoplasms of the uterus, ovary, and skin. While the associations with reproductive tract neoplasms may arise due to incidental findings during diagnostic procedures, it cannot be ruled out that some women have a genetic predisposition to tissue overgrowth that may manifest in either benign or malignant neoplasms in certain tissues. Apart from a few small-scale reports (Unler *et al*., 2016), there are no relevant studies evaluating the prevalence of different neoplasms among women with FGT polyps, therefore, our observations need to be confirmed and further evaluated in future studies.

We further saw significant associations of FGT polyps with uterine leiomyoma and endometriosis. Though the cellular composition, characteristics, and mechanism of origin of uterine fibroids and endometrial polyps differ from each other, they can occur concurrently due to common risk factors like advanced age, obesity, and hormonal dysregulation (Kınay *et al*., 2016; Pavone *et al*., 2018). Further, increased prevalence of endometriosis among women with FGT polyps emphasises the reported evidence of a higher prevalence of endometrial polyps in endometriosis patients (Shen *et al*., 2011; Zhang *et al*., 2018; Lin *et al*., 2020; Peters *et al*., 2022).

Our genetic correlation analysis showed a significant negative correlation between SHBG levels and the risk of FGT polyps. In females, testosterone and oestrogen levels are regulated through their binding to SHBG, which helps maintain hormonal balance in the bloodstream. Lower levels of SHBG can result in elevated levels of free testosterone and free oestrogen. As EPs are oestrogen dependent, lower SHBG may directly stimulate EP development. Moreover, the expression of aromatase 450 enzyme in the EPs, converts free testosterone into oestrogen, further augments the stimulation of polyps’ development (Filho *et al*., 2007). Thus, our study’s finding of the negative correlation of SHBG in FGT polyps concords with this mechanism. Unexpectedly, our analysis did not reveal a genetic correlation between oestradiol and EPs. The discrepancy between circulating plasma levels and localized endometrial levels of oestradiol may underlie the lack of this correlation. In the literature, serum levels of oestradiol were shown to be significantly lower (five times) than the endometrial oestradiol concentration in the proliferative phase (Huhtinen *et al*., 2012). The same trend was observed in women with abnormal uterine bleeding and hyperplasia (Cortés-Gallegos *et al*., 1975). Given that abnormal bleeding and hyperplasia are important symptoms of endometrial polyps, it suggests that localised oestradiol may play a more crucial role in EP development. Additionally, a study reported no significant difference in circulating oestradiol concentration among women with and without endometrial polyps (Cortés-Gallegos *et al*., 1975), which further supports our finding. This can also explain the lack of correlation of FGT polyps with BMI to some extent, as higher BMI is often associated with increased circulating oestradiol levels.

To understand how genetic variation impacts trait susceptibility, it is important to link associated genetic variants with specific genes and mechanisms. We employed a diverse range of data layers to map potential candidate genes for the identified genomic loci. Among others, *PRIM1* (rs2277339) and *COL17A1* (rs193097753) were prioritised since the GWAS signals tagged coding variants in these genes. In the literature, neither of these genes has been directly correlated with FGT development. The *PRIM1* gene is a key regulator of DNA replication during cellular proliferation process. It has been shown that the *PRIM1* expression was upregulated in breast tumour tissues compared to healthy tissues, and its inhibition led to tumour cell growth regression (Lee *et al*., 2019). Furthermore, the *PRIM1*-induced tumour cell growth is stimulated by oestrogen through activation of the oestrogen receptor (ER) (Lee *et al*., 2019). As per one of the postulated hypotheses, polyp development is related to oestrogen stimulation with predominantly increased ER alpha (ER-alpha) and decreased progesterone receptors (PRs) A and B in the glandular epithelium. On the other hand, in the stromal cells of polyps, lower concentrations of ER and PR hinder the decidualization process, preventing them from shedding off during menstruation (Mittal *et al*., 1996; Peng *et al*., 2009; Nijkang *et al*., 2019). Since the *PRIM1* gene signalling is oestrogen-associated, this can provide important insights into the probable functional mechanisms underlying the development of polyps. The *COL17A1* gene is closely associated with epithelial tumour progression and invasiveness (Jones *et al*., 2020), suggesting that a shared biological mechanism related to epithelial cell proliferation could be associated with the development of EPs. However, further functional assays on *PRIM1* and *COL17A1* genes are needed to confirm these associations.

While our study is a large-scale study, it had certain limitations. The cohort from Estonian and Finnish biobanks encompasses women with all kinds of reproductive polyps, and our phenotype definition did not differentiate between the types/location of polyps to increase study sample size power. However, considering the overall occurrence of polyps in the general population, endometrial polyps would be more prevalent than the other types. A complete understanding of the transcriptome profile of endometrial polyps is still lacking, resulting in knowledge gaps regarding the interplay between genomic loci and the regulation of gene expression. By addressing these knowledge gaps, we can gain valuable insights into the genetic factors and gene expression patterns that contribute to the pathogenesis of endometrial polyps and potentially identify novel therapeutic targets for their management.

In conclusion, the first GWAS meta-analysis of FGT polyps highlights and clarifies the genetic mechanisms shared between EP development (tissue overgrowth) and cancerous processes. Furthermore, an analysis of associated diagnoses showed that women diagnosed with EPs have an increased prevalence of other diagnoses, such as endometriosis, uterine fibroids and both benign and malignant neoplasms, which could have implications for patient management and counselling.

## Supplementary data

Supplementary data are available at Human reproduction online.

## Data availability

FinnGen cohort level summary statistics can be accessed as described here: https://www.finngen.fi/en/access_results. Protocol for accessing the Estonian Biobank data is described here: https://genomics.ut.ee/en/content/estonian-biobank. GWAS meta-analysis summary statistics will be made available via the GWAS Catalogue (data upload pending).

## Author’s roles

Authors A.D.S.P., N.P.G, A.S., M.P and T.L. participated in the conception of the study, data analysis, and interpretation of the data, and writing the manuscript. V.R., J.D., and R.M. contributed to data analysis and interpretation. T.L., M.S., M.P., V.R., J.D. and A.S. critically reviewed and provided feedback on each version of the manuscript and all the authors approved the final version. The EstBB Research Team provided the EstBB genotype and phenotype data.

## Acknowledgements

We want to acknowledge the participants of the Estonian Biobank and the participants and investigators of the FinnGen study. We also acknowledge the Estonian Biobank Research team members Andres Metspalu, Tonu Esko, Mari Nelis, Georgi Hudjashov, and Lili Milani.

## Supplementary materials

**Supplementary figure 1: Genome-wide gene association analysis of our GWAS dataset using ‘Multi-marker Analysis of GenoMic Annotation (MAGMA) wherein input SNPs were mapped to 18895 protein-coding genes.** On the Manhattan plot, the y-axis represents−log10(P-values) for the association of variants identified in polyps of the female genital tract. The horizontal red dashed line represents the genome-wide significance threshold (P < 5 × 10−8) and prioritised genes for each locus are shown. Genes associated with exonic missense variants are denoted in red text.

## Supplementary Tables

ST 1 Genes mapped by expression quantitative trait loci (eQTL) and Chromatin interactions by Functional Mapping and Annotation of Genome-Wide Association Studies (FUMA GWAS)

ST 2 Results of the Multi-marker Analysis of GenoMic Annotation (MAGMA) gene-based analysis

ST 3 Results of the GWAS catalogue lookup

ST 4 Look-up of the 10 lead variants in endometrial cancer summary statistics (https://pubmed.ncbi.nlm.nih.gov/31040137/)

ST 5 Genetic correlation between female genital tract polyps and other traits ST 6 Phenotypes associated with Polyps of the female genital tract

ST 7 Results of survival analysis lookup in FinnGen data

## References

Altaner S, Gucer F, Tokatli F, Guresci S, Ozdemir C, Puyan FO, Kutlu K. Expression of Bcl-2 and Ki-67 in tamoxifen-associated endometrial polyps: comparison with postmenopausal polyps. Onkologie 2006;29:376–380.

Annan JJ, Aquilina J, Ball E. The management of endometrial polyps in the 21st century. The Obstetrician & Gynaecologist 2012;14:33–38.

Antunes A, Costa-Paiva L, Arthuso M, Costa JV, Pinto-Neto AM. Endometrial polyps in pre- and postmenopausal women: factors associated with malignancy. Maturitas 2007;57:415–421.

Azevedo JM da R de, Azevedo LM da R de, Freitas F, Wender MCO. Endometrial polyps: when to resect? Arch Gynecol Obstet 2016;293:639–643.

Banas T, Pitynski K, Mikos M, Cielecka-Kuszyk J. Endometrial Polyps and Benign Endometrial Hyperplasia Have Increased Prevalence of DNA Fragmentation Factors 40 and 45 (DFF40 and DFF45) Together With the Antiapoptotic B-Cell Lymphoma (Bcl-2) Protein Compared With Normal Human Endometria. Int J Gynecol Pathol 2018;37:431–440.

Bateman NW, Dubil EA, Wang G, Hood BL, Oliver JM, Litzi TA, Gist GD, Mitchell DA, Blanton B, Phippen NT, et al. Race-specific molecular alterations correlate with differential outcomes for black and white endometrioid endometrial cancer patients. Cancer 2017;123:4004–4012.

Bettocchi S, Achilarre M, Ceci O, Luigi S. Fertility-Enhancing Hysteroscopic Surgery. Semin Reprod Med 2011;29:075–082.

Boyle AP, Hong EL, Hariharan M, Cheng Y, Schaub MA, Kasowski M, Karczewski KJ, Park J, Hitz BC, Weng S, et al. Annotation of functional variation in personal genomes using RegulomeDB. Genome Res 2012;22:1790–1797.

Cha P-C, Takahashi A, Hosono N, Low S-K, Kamatani N, Kubo M, Nakamura Y. A genome-wide association study identifies three loci associated with susceptibility to uterine fibroids. Nat Genet 2011;43:447–450.

Chen X, Chen S, Li Y, Gao Y, Huang S, Li H, Zhu Y. SMURF1-mediated ubiquitination of ARHGAP26 promotes ovarian cancer cell invasion and migration. Exp Mol Med 2019;51:1–12.

Dal Cin P, Vanni R, Marras S, Moerman P, Kools P, Andria M, Valdes E, Deprest J, Van de Ven W, Van den Berghe H. Four cytogenetic subgroups can be identified in endometrial polyps. Cancer Res 1995;55:1565–1568.

Doria PLS, Moscovitz T, Tcherniakovsky M, Fernandes CE, Pompei LM, Wajman M, Nimwegen AV, Haimovich S. Association of IGF-1 CA(n) and IGFBP3 rs2854746 Polymorphisms with Endometrial Polyp Risk. Biomed Res Int 2018;2018:8704346.

Dos Santos GA, Viana NI, Pimenta R, Camargo JA de, Guimaraes VR, Romão P, Candido P, Ghazarian V, Reis ST, Leite KRM, et al. Pan-cancer analysis reveals that CTC1-STN1-TEN1 (CST) complex may have a key position in oncology. Cancer Genet 2022;262– 263:80–90.

Dreisler E, Stampe Sorensen S, Ibsen PH, Lose G. Prevalence of endometrial polyps and abnormal uterine bleeding in a Danish population aged 20-74 years. Ultrasound Obstet Gynecol 2009;33:102–108.

Edwards TL, Hartmann KE, Velez Edwards DR. Variants in BET1L and TNRC6B associate with increasing fibroid volume and fibroid type among European Americans. Hum Genet 2013;132:1361–1369.

ENCODE Project Consortium. An integrated encyclopedia of DNA elements in the human genome. Nature 2012;489:57–74.

Fatemi HM, Kasius JC, Timmermans A, Disseldorp J van, Fauser BC, Devroey P, Broekmans FJ. Prevalence of unsuspected uterine cavity abnormalities diagnosed by office hysteroscopy prior to in vitro fertilization. Hum Reprod 2010;25:1959–1965.

Filho AMB de B, Barbosa IC, Maia H, Genes CC, Coutinho EM. Effects of subdermal implants of estradiol and testosterone on the endometrium of postmenopausal women. Gynecol Endocrinol 2007;23:511–517.

Fung JN, Mortlock S, Girling JE, Holdsworth-Carson SJ, Teh WT, Zhu Z, Lukowski SW, McKinnon BD, McRae A, Yang J, et al. Genetic regulation of disease risk and endometrial gene expression highlights potential target genes for endometriosis and polycystic ovarian syndrome. Sci Rep 2018;8:11424.

Ghoussaini M, Mountjoy E, Carmona M, Peat G, Schmidt EM, Hercules A, Fumis L, Miranda A, Carvalho-Silva D, Buniello A, et al. Open Targets Genetics: systematic identification of trait-associated genes using large-scale genetics and functional genomics. Nucleic Acids Res 2021;49:D1311–D1320.

GTEx Consortium. The Genotype-Tissue Expression (GTEx) project. Nat Genet 2013;45:580– 585.

Hinckley MD, Milki AA. 1000 office-based hysteroscopies prior to in vitro fertilization: feasibility and findings. JSLS 2004;8:103–107.

Humaidan P, Van Vaerenbergh I, Bourgain C, Alsbjerg B, Blockeel C, Schuit F, Van Lommel L, Devroey P, Fatemi H. Endometrial gene expression in the early luteal phase is impacted by mode of triggering final oocyte maturation in recFSH stimulated and GnRH antagonist co-treated IVF cycles. Hum Reprod 2012;27:3259–3272.

Jones VA, Patel PM, Gibson FT, Cordova A, Amber KT. The Role of Collagen XVII in Cancer: Squamous Cell Carcinoma and Beyond. Front Oncol 2020;10:352.

Karayalcin R, Ozcan S, Moraloglu O, Ozyer S, Mollamahmutoglu L, Batıoglu S. Results of 2500 office-based diagnostic hysteroscopies before IVF. Reprod Biomed Online 2010;20:689–693.

Kho PF, Wang X, Cuéllar-Partida G, Dörk T, Goode EL, Lambrechts D, Scott RJ, Spurdle AB, O’Mara TA, Glubb DM. Multi-tissue transcriptome-wide association study identifies eight candidate genes and tissue-specific gene expression underlying endometrial cancer susceptibility. Commun Biol 2021;4:1211.

Kircher M, Witten DM, Jain P, O’Roak BJ, Cooper GM, Shendure J. A general framework for estimating the relative pathogenicity of human genetic variants. Nat Genet 2014;46:310–315.

Kınay T, Öztürk Başarır Z, Fırtına Tuncer S, Akpınar F, Kayıkçıoğlu F, Koç S. Prevalence of endometrial polyps coexisting with uterine fibroids and associated factors. Turk J Obstet Gynecol 2016;13:31–36.

Koel M, Võsa U, Jõeloo M, Läll K, Gualdo NP, Laivuori H, Lemmelä S; Estonian Biobank Research Team; FinnGen; Daly M, et al. GWAS meta-analyses clarify the genetics of cervical phenotypes and inform risk stratification for cervical cancer. Hum Mol Genet. 2023,32:2103–2116.

Kurki MI, Karjalainen J, Palta P, Sipilä TP, Kristiansson K, Donner KM, Reeve MP, Laivuori H, Aavikko M, Kaunisto MA, et al. FinnGen provides genetic insights from a well-phenotyped isolated population. Nature 2023;613:508–518.

Lai T, Qiu H, Si L, Zhen Y, Chu D, Guo R. Long noncoding RNA BMPR1B-AS1 facilitates endometrial cancer cell proliferation and metastasis by sponging miR-7-2-3p to modulate the DCLK1/Akt/NF-κB pathway. Cell Cycle 2022;21:1599–1618.

Laisk T, Lepamets M, Koel M, Abner E, Estonian Biobank Research Team, Mägi R. Genome-wide association study identifies five risk loci for pernicious anemia. Nat Commun 2021;12:3761.

Lee SC, Kaunitz AM, Sanchez-Ramos L, Rhatigan RM. The oncogenic potential of endometrial polyps: a systematic review and meta-analysis. Obstet Gynecol 2010;116:1197–1205.

Lee W-H, Chen L-C, Lee C-J, Huang C-C, Ho Y-S, Yang P-S, Ho C-T, Chang H-L, Lin I-H, Chang H-W, et al. DNA primase polypeptide 1 (PRIM1) involves in estrogen-induced breast cancer formation through activation of the G2/M cell cycle checkpoint. Int J Cancer 2019;144:615–630.

Leeuw CA de, Mooij JM, Heskes T, Posthuma D. MAGMA: generalized gene-set analysis of GWAS data. PLoS Comput Biol 2015;11:e1004219.

Lenci MA, Nascimento VAL do, Grandini AB, Fahmy WM, Depes D de B, Baracat FF, Lopes RGC. Premalignant and malignant lesions in endometrial polyps in patients undergoing hysteroscopic polypectomy. Einstein (Sao Paulo) 2014;12:16–21.

Liang H, Cheung LWT, Li J, Ju Z, Yu S, Stemke-Hale K, Dogruluk T, Lu Y, Liu X, Gu C, et al. Whole-exome sequencing combined with functional genomics reveals novel candidate driver cancer genes in endometrial cancer. Genome Res 2012;22:2120–2129.

Lin S, Xie X, Guo Y, Zhang H, Liu C, Yi J, Su Y, Deng Q, Zhu W. Clinical characteristics and pregnancy outcomes of infertile patients with endometriosis and endometrial polyps: A retrospective cohort study. Taiwan J Obstet Gynecol 2020;59:916–921.

Liu B, Wang T, Jiang J, Li M, Ma W, Wu H, Zhou Q. Association of BET1L and TNRC6B with uterine leiomyoma risk and its relevant clinical features in Han Chinese population. Sci Rep 2018;8:7401.

Mittal K, Schwartz L, Goswami S, Demopoulos R. Estrogen and progesterone receptor expression in endometrial polyps. Int J Gynecol Pathol 1996;15:345–348.

Nichols CA, Gibson WJ, Brown MS, Kosmicki JA, Busanovich JP, Wei H, Urbanski LM, Curimjee N, Berger AC, Gao GF, et al. Loss of heterozygosity of essential genes represents a widespread class of potential cancer vulnerabilities. Nat Commun 2020;11:2517.

Nijkang NP, Anderson L, Markham R, Manconi F. Endometrial polyps: Pathogenesis, sequelae and treatment. SAGE Open Med 2019;7:2050312119848247.

O’Mara TA, Glubb DM, Amant F, Annibali D, Ashton K, Attia J, Auer PL, Beckmann MW, Black A, Bolla MK, et al. Identification of nine new susceptibility loci for endometrial cancer. Nat Commun 2018;9:3166.

Pal L, Niklaus AL, Kim M, Pollack S, Santoro N. Heterogeneity in endometrial expression of aromatase in polyp-bearing uteri. Hum Reprod 2008;23:80–84.

Pavone D, Clemenza S, Sorbi F, Fambrini M, Petraglia F. Epidemiology and Risk Factors of Uterine Fibroids. Best Pract Res Clin Obstet Gynaecol 2018;46:3–11.

Peng X, Li T, Xia E, Xia C, Liu Y, Yu D. A comparison of oestrogen receptor and progesterone receptor expression in endometrial polyps and endometrium of premenopausal women. J Obstet Gynaecol 2009;29:340–346.

Peters M, Mikeltadze I, Karro H, Saare M, Estonian Biobank Research Team, Salumets A, Mägi R, Laisk T. Endometriosis and irritable bowel syndrome: similarities and differences in the spectrum of comorbidities. Hum Reprod 2022;37:2186–2196.

Pujol-Gualdo N, Läll K, Lepamets M, Estonian Biobank Research Team, Rossi H-R, Arffman RK, Piltonen TT, Mägi R, Laisk T. Advancing our understanding of genetic risk factors and potential personalized strategies for pelvic organ prolapse. Nat Commun 2022;13:3584.

Rahmioglu N, Mortlock S, Ghiasi M, Møller PL, Stefansdottir L, Galarneau G, Turman C, Danning R, Law MH, Sapkota Y, et al. The genetic basis of endometriosis and comorbidity with other pain and inflammatory conditions. Nat Genet 2023;55:423–436.

Raz N, Feinmesser L, Moore O, Haimovich S. Endometrial polyps: diagnosis and treatment options - a review of literature. Minim Invasive Ther Allied Technol 2021;30:278–287.

Roadmap Epigenomics Consortium, Kundaje A, Meuleman W, Ernst J, Bilenky M, Yen A, Heravi-Moussavi A, Kheradpour P, Zhang Z, Wang J, et al. Integrative analysis of 111 reference human epigenomes. Nature 2015;518:317–330.

Sahoo SS, Aguilar M, Xu Y, Lucas E, Miller V, Chen H, Zheng W, Cuevas IC, Li H-D, Hitrys D, et al. Endometrial polyps are non-neoplastic but harbor epithelial mutations in endometrial cancer drivers at low allelic frequencies. Mod Pathol 2022;35:1702–1712.

Sasaki LMP, Andrade KRC, Figueiredo ACMG, Wanderley M da S, Pereira MG. Factors Associated with Malignancy in Hysteroscopically Resected Endometrial Polyps: A Systematic Review and Meta-Analysis. J Minim Invasive Gynecol 2018;25:777–785.

Schmitt AD, Hu M, Jung I, Xu Z, Qiu Y, Tan CL, Li Y, Lin S, Lin Y, Barr CL, et al. A Compendium of Chromatin Contact Maps Reveals Spatially Active Regions in the Human Genome. Cell Rep 2016;17:2042–2059.

She J, Su D, Diao R, Wang L. A Joint Model of Random Forest and Artificial Neural Network for the Diagnosis of Endometriosis. Front Genet 2022;13:848116.

Shen L, Wang Q, Huang W, Wang Q, Yuan Q, Huang Y, Lei H. High prevalence of endometrial polyps in endometriosis-associated infertility. Fertil Steril 2011;95:2722–2724.e1.

Shiratori A, Okumura K, Nogami M, Taguchi H, Onozaki T, Inoue T, Ando T, Shibata T, Izumi M, Miyazawa H. Assignment of the 49-kDa (PRIM1) and 58-kDa (PRIM2A and PRIM2B) subunit genes of the human DNA primase to chromosome bands 1q44 and 6p11.1-p12. Genomics 1995;28:350–353.

Takeda T, Banno K, Kobayashi Y, Adachi M, Yanokura M, Tominaga E, Kosaki K, Aoki D. Mutations of RAS genes in endometrial polyps. Oncol Rep 2019;42:2303–2308.

Tanos V, Berry KE, Seikkula J, Abi Raad E, Stavroulis A, Sleiman Z, Campo R, Gordts S. The management of polyps in female reproductive organs. Int J Surg 2017;43:7–16.

Uglietti A, Buggio L, Farella M, Chiaffarino F, Dridi D, Vercellini P, Parazzini F. The risk of malignancy in uterine polyps: A systematic review and meta-analysis. Eur J Obstet Gynecol Reprod Biol 2019;237:48–56.

Unler GK, Gokturk HS, Toprak E, Erinanc OH, Korkmaz H. Does the Presence of Endometrial Polyp Predict Colorectal Polyp? Am J Med Sci 2016;351:129–132.

Vieira MDC, Vitagliano A, Rossette MC, Neto LC de A, Gallo A, Sardo ADS. Endometrial Polyps: Update Overview on Etiology, Diagnosis, Natural History and Treatment. CEOG 2022;49:232. IMR Press.

Vitale SG, Haimovich S, Laganà AS, Alonso L, Di Spiezio Sardo A, Carugno J, From the Global Community of Hysteroscopy Guidelines Committee. Endometrial polyps. An evidence-based diagnosis and management guide. Eur J Obstet Gynecol Reprod Biol 2021;260:70–77.

Wang K, Li M, Hakonarson H. ANNOVAR: functional annotation of genetic variants from high-throughput sequencing data. Nucleic Acids Res 2010;38:e164.

Watanabe K, Taskesen E, Bochoven A van, Posthuma D. Functional mapping and annotation of genetic associations with FUMA. Nat Commun 2017;8:1826.

Wu L, Wang Y, Liu Y, Yu S, Xie H, Shi X, Qin S, Ma F, Tan TZ, Thiery JP, et al. A central role for TRPS1 in the control of cell cycle and cancer development. Oncotarget 2014;5:7677–7690.

Yi SE, LaPolt PS, Yoon BS, Chen JY, Lu JK, Lyons KM. The type I BMP receptor BmprIB is essential for female reproductive function. Proc Natl Acad Sci U S A 2001;98:7994– 7999.

Zhang Y-N, Zhang Y-S, Yu Q, Guo Z-Z, Ma J-L, Yan L. Higher Prevalence of Endometrial Polyps in Infertile Patients with Endometriosis. Gynecol Obstet Invest 2018;83:558– 563.

