## Supplementary material for "Large-scale genome-wide association study to determine the genetic underpinnings of female genital tract polyps": Supplementary Figure 1.docx

Supplementary Figure 1. **Genome-wide gene association analysis of our GWAS dataset using ‘Multi-marker Analysis of GenoMic Annotation (MAGMA) wherein input SNPs were mapped to 18895 protein-coding genes.**

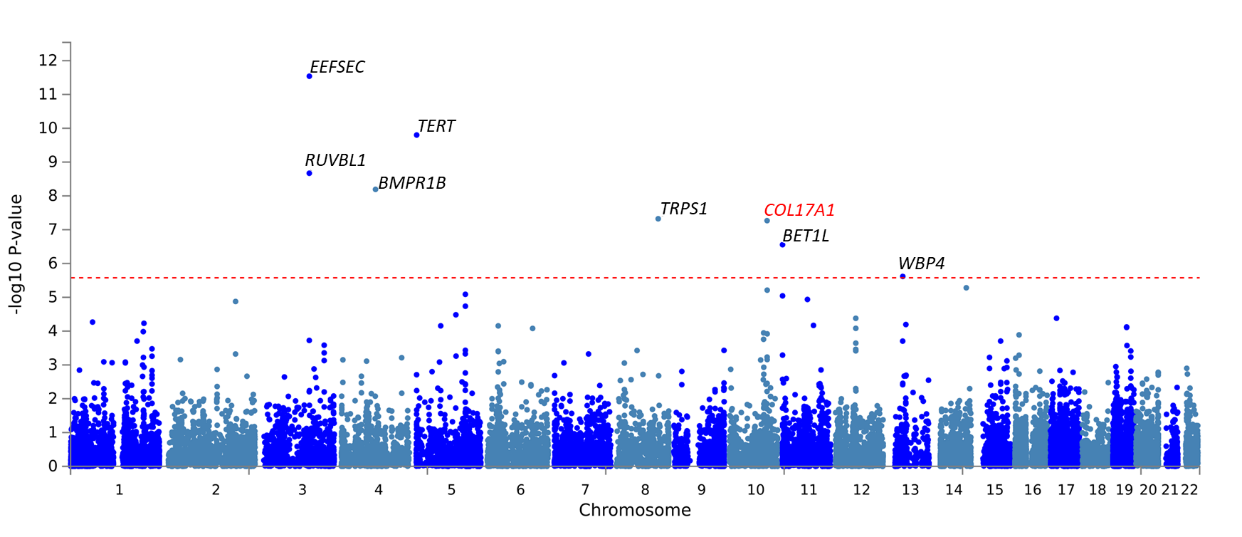

On the Manhattan plot, the y-axis represents −log10(P-values) for the association of variants identified in polyps of the female genital tract. The horizontal red dashed line represents the genome-wide significance threshold (P < 5 × 10^−8^) and prioritised genes for each locus are shown. Genes associated with exonic missense variants are denoted in red text.
